# Detecting PI3K and TP53 Pathway Disruptions in Early-Onset Colorectal Cancer Among Hispanic/Latino Patients

**DOI:** 10.1101/2024.11.13.24317265

**Authors:** Cecilia Monge, Brigette Waldrup, Sophia Manjarrez, Francisco Carranza, Enrique Velazquez-Villarreal

## Abstract

**Background/Objectives:** This study aims to characterize PI3K and TP53 pathway alterations in Hispanic/Latino patients with early-onset colorectal cancer (CRC), focusing on potential differences compared to non-Hispanic White (NHW) patients. Understanding these differences may shed light on the molecular basis of CRC health disparities.

**Methods:** Using cBioPortal, we conducted a bioinformatics analysis to evaluate CRC mutations within the PI3K and TP53 pathways. CRC cases were stratified by age and ethnicity: (1) early-onset (<50 years) versus late-onset (≥50 years) and (2) early-onset in Hispanic/Latino patients compared to early-onset in NHW patients. Mutation frequencies were assessed using descriptive statistics, with chi-squared tests comparing proportions between early-onset Hispanic/Latino and NHW groups. Kaplan-Meier survival curves were generated to assess overall survival for early-onset Hispanic/Latino patients, stratified by the presence or absence of PI3K and TP53 pathway alterations.

**Results:** Significant differences were noted when comparing early-onset CRC in Hispanic/Latino patients to early-onset CRC in NHW patients. PI3K pathway alterations were more prevalent in early-onset CRC among Hispanic/Latino patients (90.5% vs. 41.5%, p = 9.279e-5), with mTOR alterations also significantly higher in this group (14.3% vs. 1.5%, p = 0.043). No significant differences were observed between early-onset and late-onset CRC cases within the Hispanic/Latino cohort. Additionally, Hispanic/Latino patients with early-onset CRC showed improved clinical outcomes when TP53 pathway alterations were present (p =5.4e-4).

**Conclusions:** These findings highlight the distinct role of PI3K and TP53 pathway disruptions in early-onset CRC among Hispanic/Latino patients, suggesting that pathway-specific mechanisms may drive cancer health disparities. Insights from this study could inform the potential development of precision medicine approaches and targeted therapies aimed at addressing these disparities and improving outcomes for diverse patient populations.

## 1. Introduction

Colorectal Cancer (CRC) is third most prevalent cancer type and the second most common cancer-related cause of death globally (1). While the overall incidence of colorectal cancer has stabilized or decreased in higher-income countries, a concerning trend has emerged, showing a rise in CRC among individuals under 50 years old (2, 3). This disturbing trend is also accompanied by an increase in colorectal cancer-related mortality (4). This trend is further exacerbated when viewed through the lens of health disparities in the Hispanic/Latino (H/L) population. The H/L population has the highest increase in early-onset CRC (EOCRC) incidence when compared to all other races in the US (5). The H/L population also has the highest increase in EOCRC mortality when compared to their non-Hispanic white counterparts (6, 7). To effectively address this significant public health concern, several factors must be explored, including genomic determinants.

EOCRC is often diagnosed at more advanced stages, possibly because preventive screening measures typically do not begin until the age of 50 (8). However, studies have emerged revealing EOCRC has distinct molecular characteristics such as significantly higher microsatellite instability distribution rate, elevated tumor mutation burden and PD-L1 expression (2, 9). In addition, LINE-1 hypomethylation has been suggests a distinct molecular biomarker of EOCRC (10). Multiple signaling pathways are implicated in the development and progression of CRC. Two key pathways involved are the PI3K and TP53 pathways.

PI3K pathway is a primary signaling pathway that regulates processes, including cell metabolism, apoptosis and proliferation (11). The pathway is controlled by four principal types of receptors: receptor tyrosine kinases (RTKs), which sense growth factors; cytokine receptors, associated with immune responses and cell growth; G-protein coupled receptors (GPCRs), linked to metabolic functions; and integrins, which play a role in sensing cell-cell and cell-matrix adhesion. Each of these receptor type initiates signaling cascades that regulate essential cellular processes, such as survival, growth, and communication (12). Hyperactivation of the PI3K pathway has been associated with tumor cell proliferation, cell invasion, and reduced apoptosis (13). In CRC, the most common genetic alterations include IGF2 overexpression, PIK3CA mutations, and PTEN mutations or deletions, with these changes present in approximately 40% of malignant tumors (12). Clinical data has shown advanced stages of CRC with mutations in this pathway do not respond well to anti EGFR therapy (14). Other key genes altered in this pathway that contribute to CRC development include AKT1 mutations, which are associated with disease progression and resistance to therapy via various downstream targets (12, 15). mTOR complex 1 (mTORC1), which is activated by fully phosphorylated AKT, is one of the key targets within this signaling pathway (12). mTORC1 plays a crucial role in regulating many metabolic processes that contribute to tumor growth and drug resistance. One example is its role as a key contributor to drug resistance in treatments targeting PI3K inhibitors (15). Although alterations in the PI3K/AKT pathway play a critical role in CRC development, these aberrations have not been well-characterized in EOCRC in the H/L population.

The TP53 pathway regulates a wide range of cellular functions, including cell cycle arrest, DNA repair, apoptosis, autophagy, and metabolism. The primary gene in the TP53 pathway is the transcription factor p53, which located in the nucleus and cytoplasm and specifically binds to DNA to regulate a diverse set of genes. P53, also known as the guardian of the genome, plays a crucial role in maintain genomic stability and coordinating DNA damage repair response mechanisms and promoting cell-cycle arrest through the p21 and repressing cyclin-dependent kinases (CDKs) and cyclin B (16). CRC exhibits one of the highest incidences of TP53 mutations, with approximately 74% of tumors harboring a TP53 mutation. Currently, numerous drugs are undergoing clinical trials aimed at validating their safety and efficacy for use in CRC treatment (17). Additionally, gain-of-function missense mutations in TP53 have been shown to contribute to therapy resistance (18). Although alterations in the TP53 pathway are crucial for CRC development, their specific role in EOCRC, especially among Latino populations, remains poorly understood.

In this study, we aim to perform a comprehensive molecular analysis of the PI3K and TP53 signaling pathways in colorectal cancer within the H/L population, comparing early-onset CRC cases with those diagnosed in H/L patients over the age of 50. We also investigated other molecular characteristics of early-onset colorectal cancer, including tumor mutation burden and common driver oncogenes seen in CRC in H/L patients.

## 2. Materials and Methods

To conduct our analysis, we utilized clinical and genomic data from 19 colorectal cancer (CRC) datasets accessed via the cBioPortal database. These included studies classified under colorectal adenocarcinoma, colon adenocarcinoma, and rectal adenocarcinoma, as well as data from the GENIE BPC CRC v2.0-public dataset. Two studies focusing on metastatic CRC were excluded from this analysis. Following dataset selection, we applied a series of filtering criteria to refine our sample pool. The criteria included selecting patients identified as Hispanic or Latino, Spanish, NOS; Hispanic, NOS; Latino, NOS; or those with a Mexican or Spanish surname; restricting samples to primary tumor cases; including only colon adenocarcinoma, rectal adenocarcinoma, and colorectal adenocarcinoma; confirming histology as adenocarcinoma, NOS; and ensuring one sample per patient. This process resulted in three datasets meeting all criteria: TCGA PanCancer Atlas, MSK Nat Commun 2022, and GENIE BPC CRC, comprising 20 early-onset and 13 late-onset CRC samples from Hispanic/Latino patients. Age at diagnosis was extracted from individual clinical records within the GENIE database.

Pathway alterations were defined according to previously established criteria (18). To create our analysis cohorts, we categorized patients into early-onset (under 50 years of age) and late-onset (50 years or older) groups. Ethnicity-based classification divided participants into Hispanic/Latino and Non-Hispanic White (NHW) groups. We further stratified these groups based on the presence or absence of PI3K and TP53 pathway alterations to enable a detailed examination of the interactions between age, ethnicity, and these molecular changes.

Statistical analysis included Chi-square tests to evaluate the independence of categorical variables and identify potential associations between age, ethnicity, and pathway alterations. Additionally, we stratified samples by tumor location, distinguishing between colon and rectal cancers. This level of stratification facilitated a nuanced analysis of the interplay between age, ethnicity, and tumor location in relation to pathway disruptions, offering deeper insights into patient heterogeneity and potential implications for treatment responses.

Kaplan-Meier survival analysis was employed to assess overall survival, focusing on the impact of PI3K and TP53 pathway alterations. Survival curves were constructed to illustrate survival probabilities over time, with patients grouped by the presence or absence of pathway disruptions. The log-rank test was utilized to determine statistically significant differences between survival curves. Median survival times were calculated, accompanied by 95% confidence intervals to convey the precision of these estimates. This comprehensive methodological approach provided an in-depth understanding of how specific pathway alterations may affect patient outcomes in early-and late-onset CRC cases within the Hispanic/Latino population.

## 3. Results

From three cBioPortal projects that reported ethnicity, we constructed our Hispanic/Latino (H/L) cohort, which comprised 33 samples, while the Non-Hispanic White (NHW) cohort included 341 samples. In the H/L cohort, 63.7% of patients presented with early-onset colorectal cancer (CRC) (diagnosed before age 50), while 36.3% were diagnosed at age 50 or older (Table 1). Comparatively, in the NHW cohort, 19% had early-onset CRC, with 81% being diagnosed at age 50 or older. The H/L cohort consisted of 63.6% male and 36.4% female patients, whereas the NHW cohort was composed of 53.5% male and 46.5% female patients. At the time of diagnosis, 15.2% of patients in the H/L cohort were at an early stage (Stages 0, I, and II), while 69.7% were at a late stage (Stage III or IV). In contrast, the NHW cohort had 4.39% of patients diagnosed at an early stage and 10.5% at a late stage, with 85% of NHW patients recorded as NA for stage at diagnosis. All patients in the H/L cohort identified as Hispanic or Latino, which highlights the targeted demographic of the study.

**Table 1.** Patient Demographics and Clinical Characteristics of the Hispanic/Latino (H/L) and Non-Hispanic White (NHW) cohorts.

| Clinical Feature | H/L Cohort<br>n (%) | NHW Cohort<br>n (%) |
| --- | --- | --- |
| **Age\* and Gender** |  |  |
| Early-Onset (< 50) Male | 12 (36.4%) | 38 (11.1%) |
| Early-Onset (< 50) Female | 9 (27.3%) | 27 (7.9%) |
| Late-Onset (≥ 50) Male | 8 (24.2%) | 144 (42.1%) |
| Late-Onset (≥ 50) Female | 4 (12.1%) | 132 (38.6%) |
| **Stage at Diagnosis\*\*** |  |  |
| Early Stage (Stage 0, I, and II) | 5 (15.2%) | 15 (4.39%) |
| Late Stage (Stage III or IV) | 23 (69.7%) | 36 (10.5%) |
| **Ethnicity** |  |  |
| Hispanic or Latino | 33 (100.0%) | 0 (0.0%) |
| Non-Hispanic or Non-Latino | 0 (0.0%) | 342 (100.0%) |

\*NHW NA:1
\*\* HL NA: 5, NHW NA: 291

The comparative analysis of clinical features between early-onset and late-onset Hispanic/Latino (H/L) colorectal cancer (CRC) patients, as well as between early-onset H/L and early-onset non-Hispanic White (NHW) patients, reveals notable distinctions (Table 2). The median age at diagnosis for early-onset H/L patients was 41 years (IQR 36-45), which was significantly younger than the median age of 63 years (IQR 58-74) observed in late-onset H/L patients (*p*<0.05). In contrast, no significant difference was detected between early-onset H/L (median 41 years) and early-onset NHW patients (median 43 years, IQR 38-47) (*p*>0.05).

**Table 2.** Ethnicity-associated differences in clinical features between Hispanic and Latino (H/L) and Non-Hispanic White (NHW) cohorts.

| Clinical Feature | Early-Onset H/L<br>n (%) | Late-Onset H/L<br>n (%) | p-value | Early-Onset H/L<br>n (%) | Early-Onset NHW<br>n (%) | p-value |
| --- | --- | --- | --- | --- | --- | --- |
| Median Age (IQR) | 41 (36-45) | 63 (58-74) | < **0.05** | 41 (36-45) | 43 (38-47) | > 0.05 |
| Median mutation count | 9 (6-10) | 24 (7-63) | > 0.05 | 9 (6-10) | 58 (7-79) | < **0.05** |
| **MTOR Mutation** |  |  |  |  |  |  |
| Present | 3 (14.3%) | 1 (8.3%) | > 0.05 | 3 (14.3%) | 1 (1.5%) | < **0.05** |
| Absent | 18 (85.7%) | 11 (91.7%) |  | 18 (85.7%) | 64 (98.5%) |  |

Mutation analysis indicated a median mutation count of 9 (IQR 6-10) for early-onset H/L patients, while late-onset H/L patients exhibited a higher, but not statistically significant, median mutation count of 24 (IQR 7-63) (p>0.05). Early-onset NHW patients, however, showed a significantly higher median mutation count of 58 (IQR 7-79) compared to early-onset H/L patients (*p*<0.05).

Furthermore, MTOR mutations were present in 14.3% of early-onset H/L patients, compared to 8.3% in late-onset H/L patients, without a significant difference between these groups (*p*>0.05). A more pronounced and statistically significant disparity was observed when comparing early-onset H/L to early-onset NHW patients, where only 1.5% of NHW cases exhibited MTOR mutations (*p*<0.05). These findings highlight potential genomic and clinical disparities between CRC subgroups that may have implications for understanding CRC pathogenesis and developing targeted interventions.

In our analysis of genetic alterations among Hispanic/Latino individuals with early-onset and late-onset colorectal cancer, we observed notable differences in the frequency of PI3K and TP53 pathway alterations (Table 3). PI3K alterations were present in 42.9% of early-onset cases compared to 75.0% of late-onset cases, although this difference did not reach statistical significance (p = 0.1451). Conversely, the absence of PI3K alterations was more frequent in early-onset cases (57.1%) than in late-onset cases (25.0%). For TP53 alterations, there was a similar high prevalence in both groups, with 90.5% of early-onset cases (19 out of 21) and 91.7% of late-onset cases showing alterations, but without statistical significance (p = 1). This indicates that TP53 mutations are consistently common across both early-onset and late-onset cases. These findings suggest that while PI3K alterations may vary more prominently between early and late-onset groups, TP53 mutations maintain a stable frequency across the age spectrum in Hispanic/Latino colorectal cancer patients.

**Table 3.**
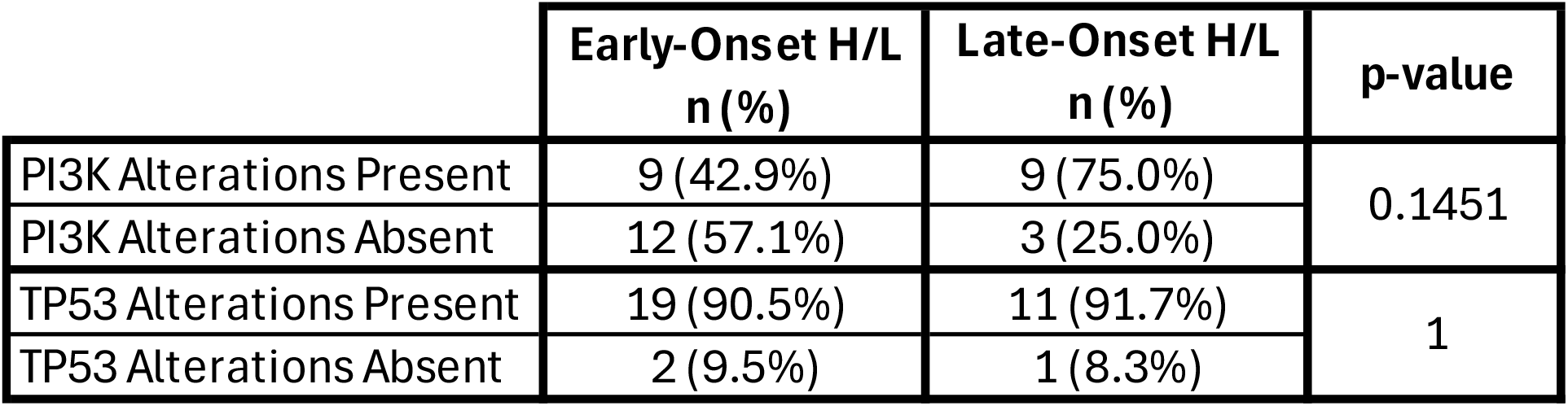
Rates of PI3K and TP53 pathway alterations among early-onset and late-onset Hispanic/Latino CRC patients.

|  | Early-Onset H/L<br>n (%) | Late-Onset H/L<br>n (%) | p-value |
| --- | --- | --- | --- |
| PI3K Alterations Present | 9 (42.9%) | 9 (75.0%) | 0.1451 |
| PI3K Alterations Absent | 12 (57.1%) | 3 (25.0%) |  |
| TP53 Alterations Present | 19 (90.5%) | 11 (91.7%) | 1 |
| TP53 Alterations Absent | 2 (9.5%) | 1 (8.3%) |  |

**Table 4.** Rates of PI3K and TP53 pathway alterations among early-onset Hispanic/Latino and Non-Hispanic White (NHW) CRC Patients.

|  | Early Onset H/L<br>n (%) | Early Onset NHW<br>n (%) | p-value |
| --- | --- | --- | --- |
| PI3K Alterations Present | 19 (90.5%) | 27 (41.5%) | 9.279E-05 |
| PI3K Alterations Absent | 2 (9.5%) | 38 (58.5%) |  |
| TP53 Alterations Present | 19 (90.5%) | 49 (75.4%) | 0.2178 |
| TP53 Alterations Absent | 2 (9.5%) | 16 (24.6%) |  |

In our analysis of genetic alterations in early-onset colorectal cancer among Hispanic/Latino (H/L) and Non-Hispanic White (NHW) individuals, we observed notable differences in the frequency of PI3K and TP53 mutations. PI3K alterations were present in 90.5% of early-onset H/L cases, compared to 41.5% of early-onset NHW. Conversely, the absence of PI3K alterations was more frequent among NHW individuals (58.5%) than among H/L individuals (9.5%). TP53 alterations, on the other hand, showed high prevalence in both early-onset groups, with 90.5% of H/L cases and 75.4% of NHW cases carrying these mutations. The absence of TP53 alterations was observed in 9.5% of H/L cases and 24.6% of NHW cases. These results highlight a potential difference in the genetic landscape between the two groups, with PI3K mutations being more common in early-onset H/L individuals than in their NHW counterparts, while TP53 alterations were consistently high across both groups. Further statistical analyses are necessary to determine the significance of these findings and explore their potential implications for precision medicine and targeted treatment approaches in colorectal cancer.

The Kaplan-Meier survival analysis for early-onset Hispanic/Latino colorectal cancer patients indicated no statistically significant difference in overall survival between those with and without PI3K pathway alterations (Figure 2). While the survival curves for patients with and without the genetic alteration appeared to diverge over time, hinting at a potential trend in survival outcomes, the p-value (p = 0.129) suggested that this observed difference was not statistically significant. The confidence intervals surrounding each curve highlighted variability in the survival estimates at different time points, emphasizing the uncertainty of these findings. These results imply that, although survival differences may exist, the current data do not provide strong evidence of a significant impact of PI3K pathway alterations on overall survival within this sample. Further research involving larger cohorts or additional stratification is needed to clarify the role of genetic alterations in survival outcomes among early-onset Hispanic/Latino colorectal cancer patients.

**Figure 1.**
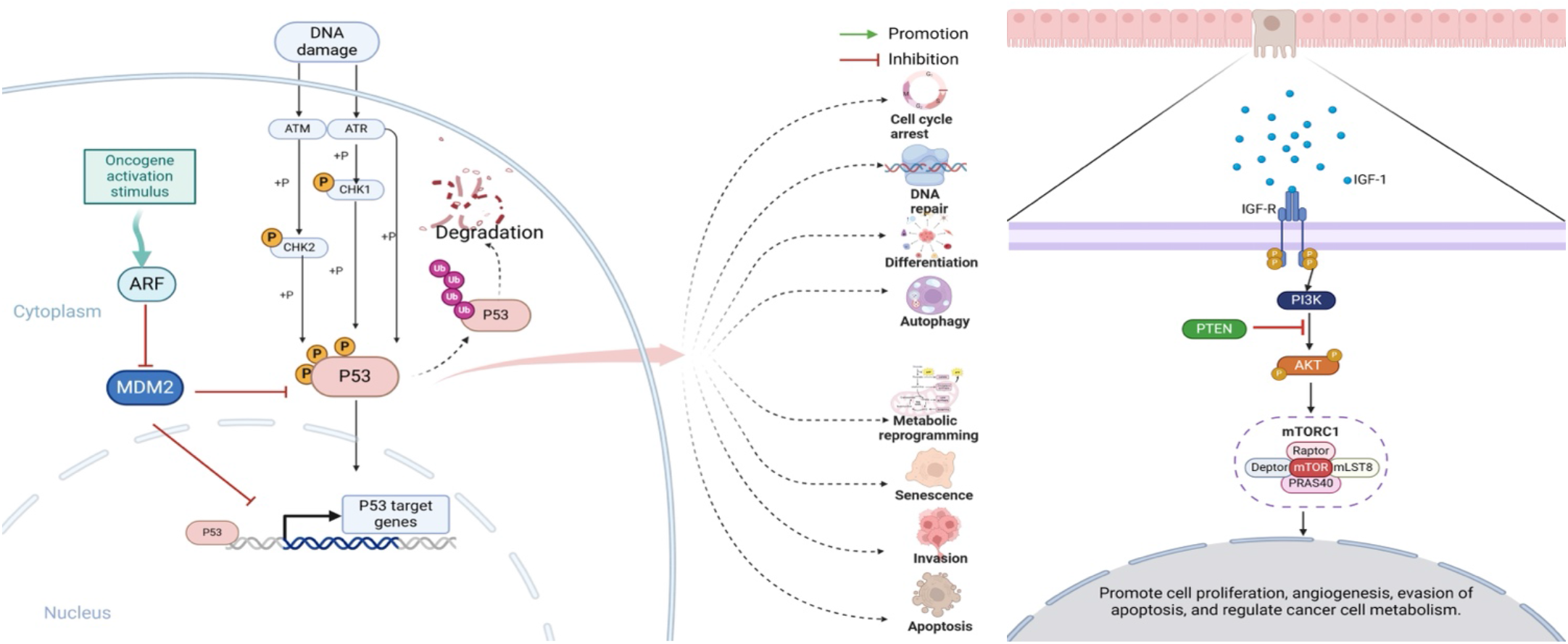
Illustration of the PI3K (left) and the and TP53 (right) signaling pathways.

**Figure 2.**
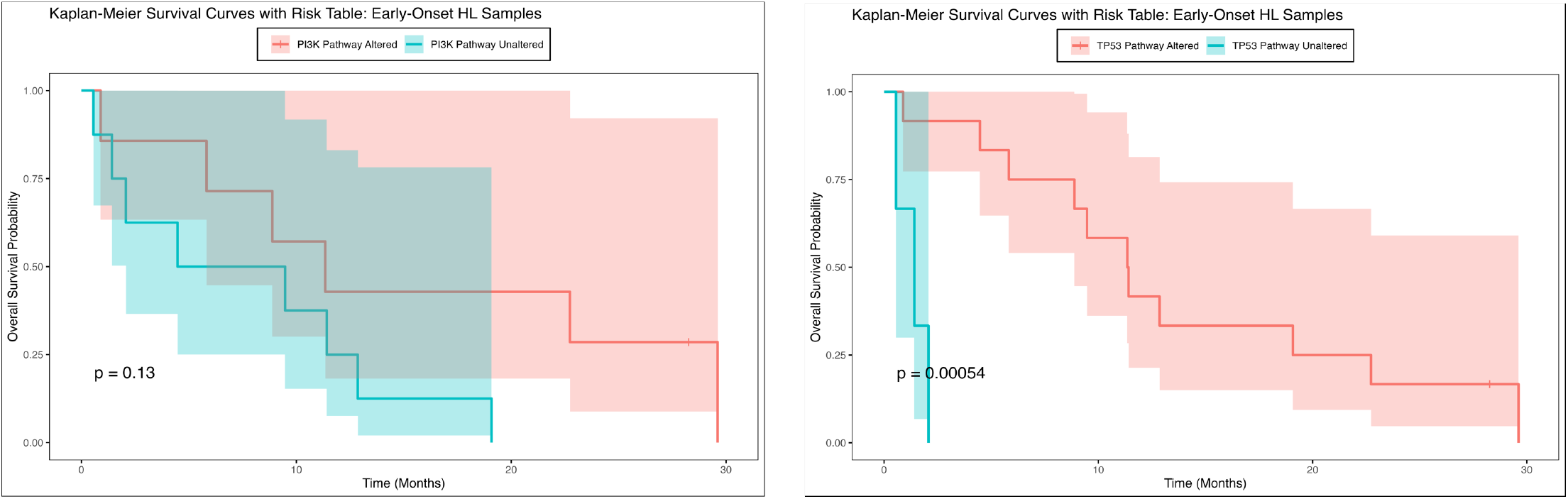
Overall survival curves of early-onset Hispanic/Latino patients stratified by the presence or absence of PI3K (left) and TP53 (right) pathway alterations.

In contrast to the findings for PI3K pathway alterations, the investigation into other genetic pathways provided more definitive insights (Figure 2). Specifically, the Kaplan-Meier survival analysis for a different subgroup of early-onset Hispanic/Latino colorectal cancer patients revealed a statistically significant difference in overall survival. Patients with the specific genetic alteration showed a marked decline in survival probability early in the follow-up period, with consistently lower survival rates compared to those without the alteration. Conversely, patients without the genetic alteration exhibited higher overall survival probabilities over time, with a more gradual decline in their survival curve. The p-value of 0.0034 underscored the statistical significance of these survival differences (p < 0.05). These findings suggest that the presence of the genetic alteration is associated with poorer prognosis and may serve as an important prognostic marker for early-onset colorectal cancer in the Hispanic/Latino population. This highlights the need for further research into targeted treatment approaches and personalized strategies to address survival disparities in this group.

Similar to the results for PI3K but in contrast to TP53 findings in the Hispanic/Latino cohort, the overall survival analysis for the NHW cohort (Figure S1) indicates that neither PI3K nor TP53 pathway alterations are significant determinants of survival outcomes in early-onset colorectal cancer within this specific ethnic group.

The alteration rates of PI3K and TP53 pathway-related genes were analyzed among early-onset and late-onset Hispanic/Latino colorectal cancer (CRC) patients to determine potential age-related differences (Table S1, Figure S2). The analysis revealed that genes associated with the PI3K pathway, such as *PIK3CA* and *PTEN*, exhibited similar alteration rates between early-onset and late-onset groups, with no statistically significant differences (e.g., *PIK3CA* alteration rates were 33% in early-onset and 58% in late-onset patients, p = 0.27). Conversely, the TP53 pathway displayed notable variations, as exemplified by *TP53*, which showed a higher alteration rate in late-onset patients (75%) compared to early-onset patients (71%), with a non-statistically significant p-value of 1. Stratification by cancer type (e.g., colon versus rectum adenocarcinoma) revealed no significant differences in PI3K and TP53 pathway alterations, indicating that these genetic variations remain consistent across CRC subtypes within this ethnic cohort (Table S2).These findings suggest that while PI3K pathway alterations may not be major determinants of CRC based on age of onset, TP53 alterations are more prevalent in late-onset cases and could play a more significant role in the disease progression of older patients. This emphasizes the need for further research to explore the implications of these genetic differences and their potential impact on targeted treatment strategies for Hispanic/Latino CRC patients across age groups.

Moreover, we compared the alteration rates of PI3K and TP53 pathway-related genes between early-onset Hispanic/Latino (H/L) and non-Hispanic White (NHW) colorectal cancer (CRC) patients (Table S3, Figure S3). The analysis revealed that while some genes, such as *TP53*, exhibited similar alteration rates between the two cohorts (71% in early-onset H/L patients versus 64% in early-onset NHW patients, p = 0.75), indicating no significant difference, other genes showed notable disparities. Specifically, mTOR alterations were more prevalent in early-onset Hispanic/Latino patients (14.3%) compared to their NHW counterparts (1.5%), with a statistically significant p-value of 0.043. When stratified by cancer type, early-onset Hispanic/Latino patients with colon adenocarcinoma did not show significant differences in PI3K and TP53 pathway alterations compared to early-onset NHW patients, suggesting that ethnic-specific variations in these pathways may not be pronounced for colon cancer (Table S4). These findings suggest that while TP53 pathway-related gene alterations may not differ substantially between these ethnic groups, PI3K pathway alterations are more pronounced in the Hispanic/Latino cohort and could be a contributing factor to ethnic disparities in CRC prognosis and outcomes. Such differences underscore the need for ethnicity-specific studies to further explore the clinical implications and inform targeted treatment strategies for early-onset CRC in diverse populations.

## 4. Discussion

The PI3K and TP53 signaling pathways are essential in the regulation of cellular functions, including cell growth, proliferation, and survival, and are frequently altered in colorectal cancer (CRC). The PI3K pathway, known for its role in cell metabolism and apoptosis, is often hyperactivated in CRC, contributing to tumorigenesis and resistance to targeted therapies [11-13]. The TP53 pathway, involving the tumor suppressor p53, regulates crucial processes such as DNA repair and cell cycle arrest and is mutated in approximately 74% of CRC cases, significantly impacting genomic stability [16-18].

While extensive research has characterized these pathways in the general population, few studies have explored their specific roles and disruption patterns in early-onset CRC among Hispanic/Latino (H/L) patients.

Our study aimed to address this knowledge gap by evaluating the alteration rates of PI3K and TP53 pathway-related genes among early-onset and late-onset CRC patients within the H/L population and comparing these findings to early-onset non-Hispanic White (NHW) patients. The results revealed significant differences in the genetic landscape of early-onset H/L CRC patients compared to their NHW counterparts. Notably, PI3K pathway alterations were more prevalent in early-onset H/L patients (90.5%) than in NHW patients (41.5%, p = 9.279e-5), with mTOR alterations significantly higher in the H/L cohort (14.3% vs. 1.5%, p = 0.043). These findings underscore the importance of understanding ethnicity-specific genetic differences, as the PI3K pathway’s hyperactivation could play a more prominent role in CRC pathogenesis in the H/L population, potentially influencing therapeutic responses and clinical outcomes.

Interestingly, our data showed no significant differences in PI3K pathway alterations when comparing early-onset to late-onset H/L CRC patients (42.9% vs. 75.0%, p = 0.1451), suggesting that PI3K-related disruptions are not strongly age-dependent in this ethnic group. Conversely, TP53 pathway alterations were consistently high in both early-onset (90.5%) and late-onset (91.7%) H/L patients (p = 1), reflecting the pathway’s critical and widespread role in CRC regardless of age at diagnosis. These results align with the well-documented role of TP53 mutations in CRC across various demographics [16-18], indicating that TP53 disruptions may be a common driver of CRC that transcends age and ethnicity.

Further, Kaplan-Meier survival analyses provided insights into the prognostic implications of these pathway alterations in early-onset H/L CRC patients. The analysis revealed no significant impact of PI3K pathway alterations on overall survival (p = 0.129), despite observed divergence in survival curves over time. In contrast, the presence of TP53 alterations was associated with poorer survival outcomes, reinforcing previous findings that highlight the tumor suppressor’s role in mediating clinical prognosis [21-24]. The consistent high prevalence of TP53 mutations across both H/L and NHW early-onset patients (90.5% and 75.4%, respectively, p = 0.75) indicates that while TP53 alterations may not vary significantly between these ethnic groups, their impact on patient outcomes remains substantial.

Our stratified analysis by cancer type (e.g., colon versus rectum adenocarcinoma) revealed no significant differences in PI3K and TP53 pathway alterations between early-onset H/L and NHW patients, suggesting that ethnic-specific variations may not be pronounced for colon cancer. However, the observed disparities in PI3K pathway alterations between the ethnic groups highlight the importance of further investigating these molecular characteristics. The higher prevalence of PI3K pathway disruptions in the H/L population could point to unique underlying biological mechanisms influenced by genetic ancestry or environmental factors [25-27]. Studies have shown that genetic backgrounds can shape CRC development, with factors such as diet and lifestyle contributing to variations in mutation profiles and disease progression [26].

The implications of these findings are significant for the development of targeted therapies and precision medicine. The elevated rates of PI3K pathway alterations in early-onset H/L patients may indicate that therapies targeting the PI3K/AKT/mTOR axis could be more effective in this population. Additionally, the consistent presence of TP53 mutations across both H/L and NHW groups underscores the need for novel strategies to target this pathway, as traditional therapeutic options remain limited [17]. High tumor mutation burden (TMB), which has been observed in early-onset CRC cases, could also be leveraged to improve treatment responses, particularly in immunotherapy settings [11].

Despite the strengths of this study, several limitations must be acknowledged. The retrospective nature of bioinformatics analyses and the potential for selection bias inherent in publicly available genomic databases may affect the generalizability of these findings. Moreover, the underrepresentation of H/L patients in genomic databases poses challenges for comprehensive analyses of molecular disparities across diverse subpopulations [25,28-30]. Larger, prospective studies are needed to confirm these findings and to explore the underlying biological mechanisms that contribute to the observed differences in pathway alterations between ethnic groups and age groups.

## 5. Conclusions

In conclusion, our findings highlight the distinct role of PI3K and TP53 pathway disruptions in early-onset CRC among Hispanic/Latino patients. The significantly higher alteration rates in the PI3K pathway and the consistent prevalence of TP53 mutations emphasize the need for ethnicity-specific studies to better understand the clinical and biological implications of these pathways. Insights from this study may inform the development of precision medicine approaches aimed at reducing CRC health disparities and improving outcomes for diverse patient populations.

## Data Availability

All data produced in the present study are available upon reasonable request to the authors and can be found online at cBioPortal.

https://www.cbioportal.org/

