## Supplemental Figures and Tables for "Detecting PI3K and TP53 Pathway Disruptions in Early-Onset Colorectal Cancer Among Hispanic/Latino Patients" (from another version of this preprint: version 2, 2024-11-15)

**Figure S1.** Overall survival curves of early-onset Non-Hispanic White (NHW) patients stratified by the presence or absence of PI3K (left) and TP53 (right) pathway alterations.

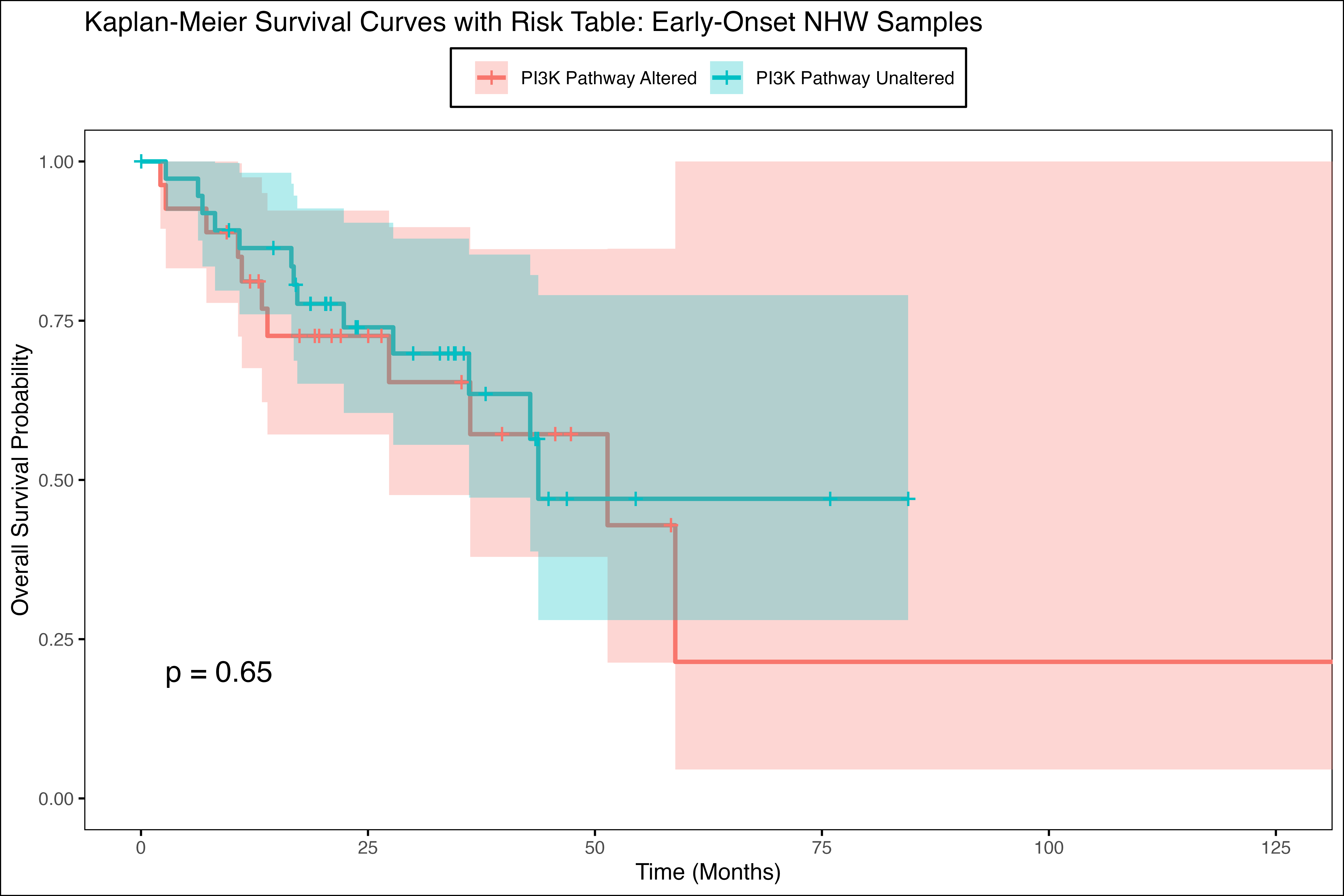

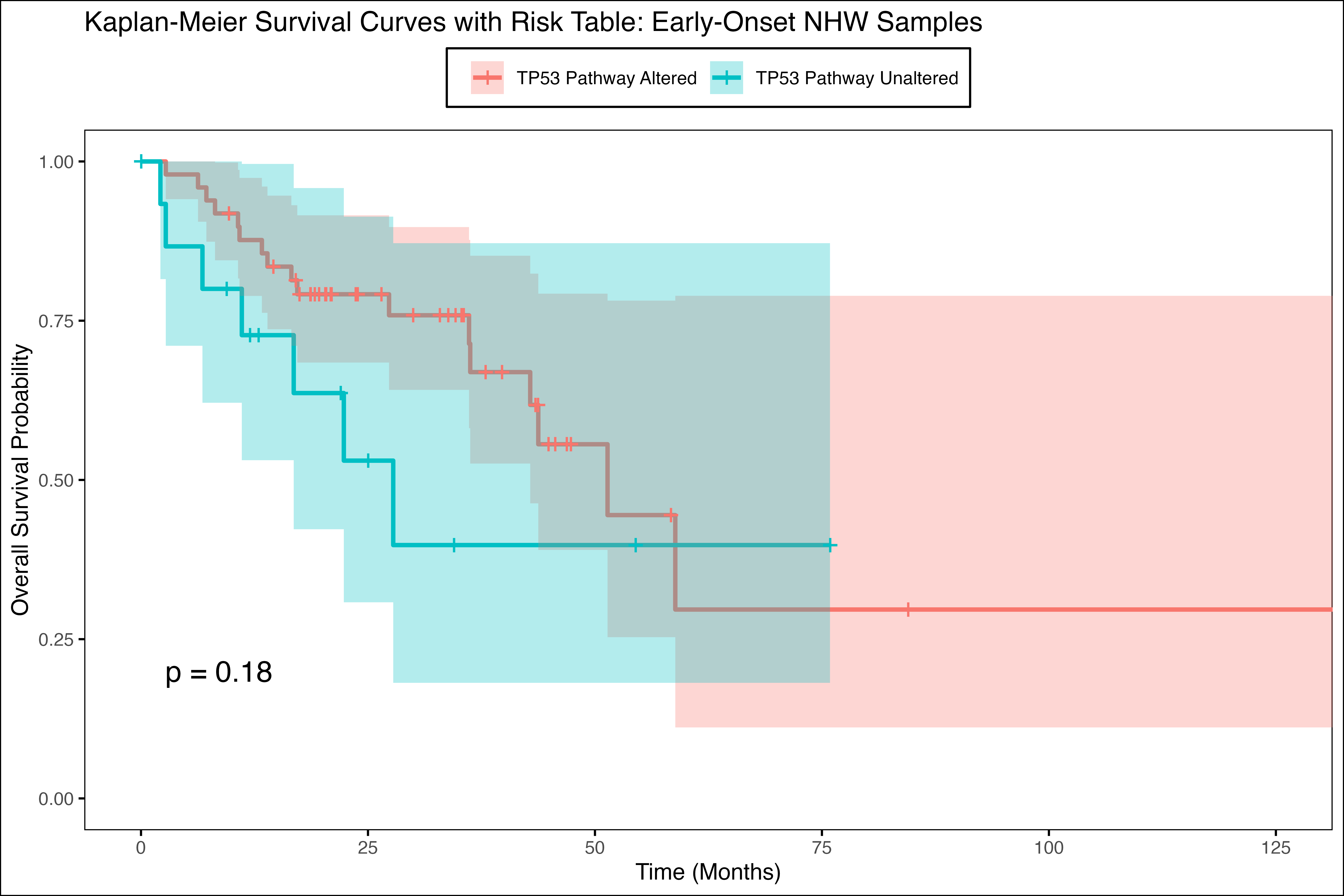

**Table S1.** Alteration Rates of PI3K and TP53 Pathway-Related Genes Among Early-Onset and Late-Onset Hispanic/Latino CRC Patients.

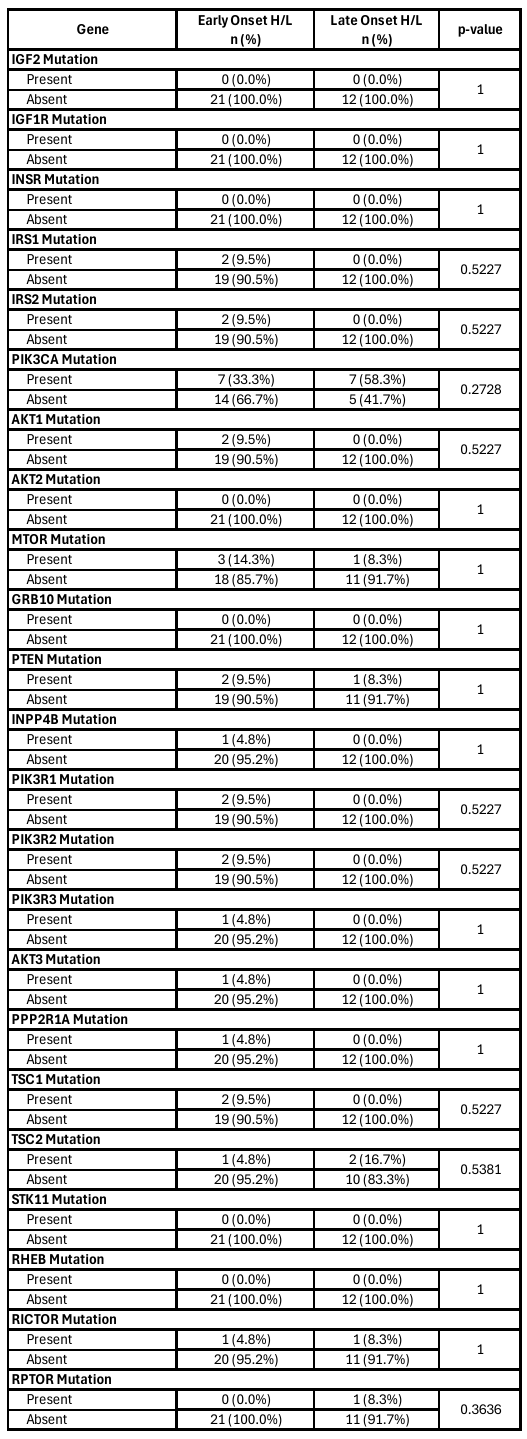

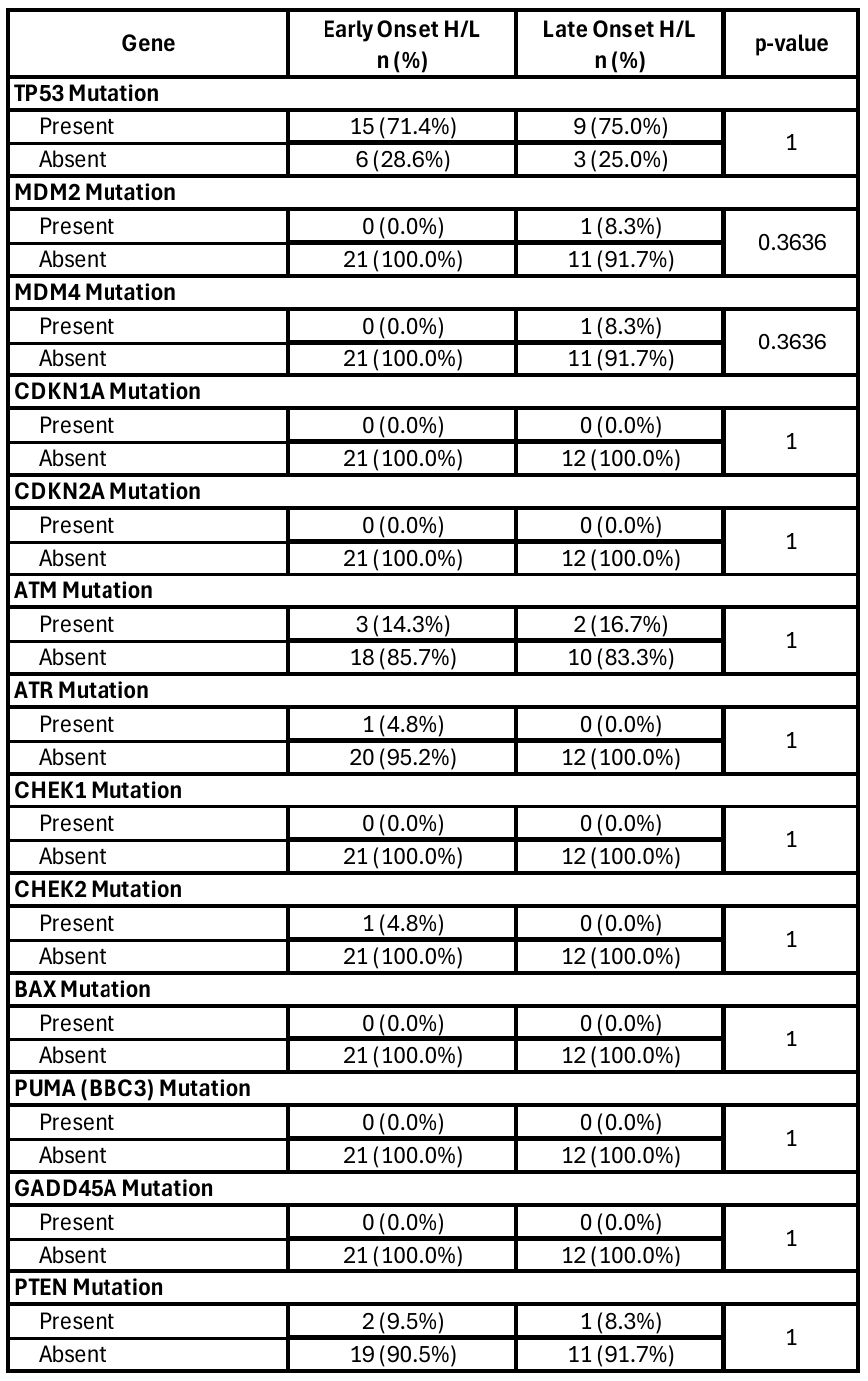

**Figure S2.** Counts of PI3K and TP53 Pathway-Related Gene Alterations in Early-Onset and Late-Onset Hispanic/Latino CRC Patients.

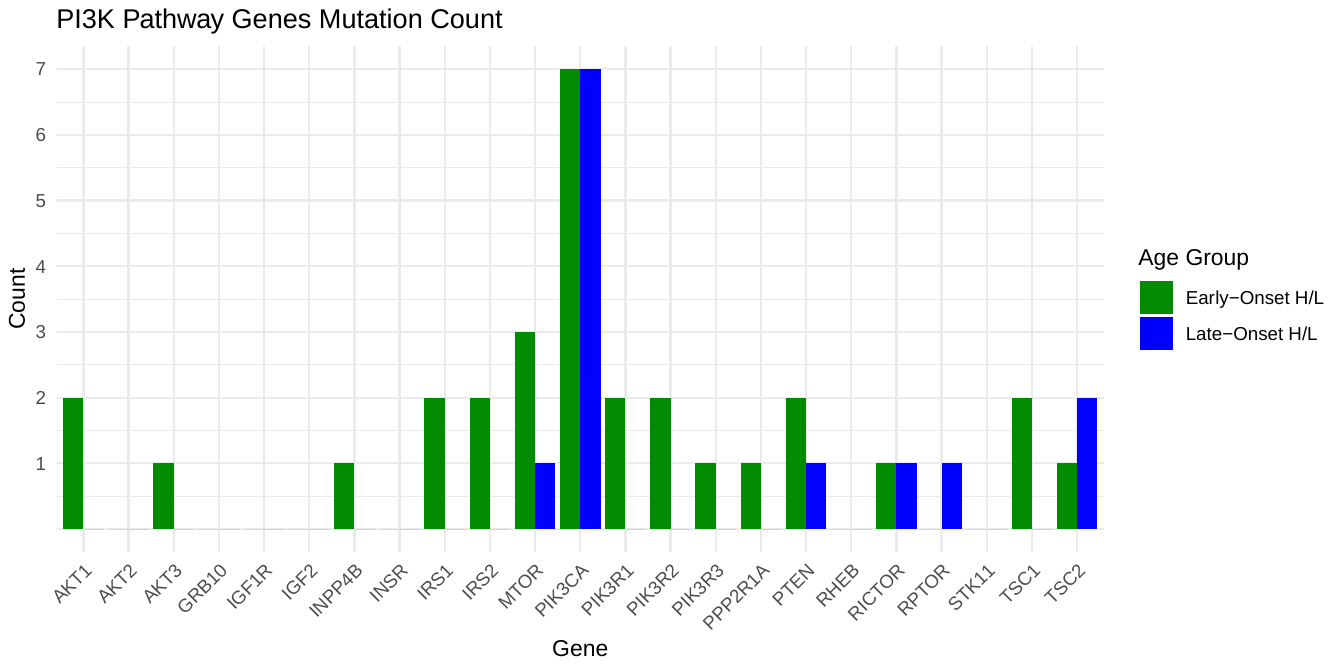

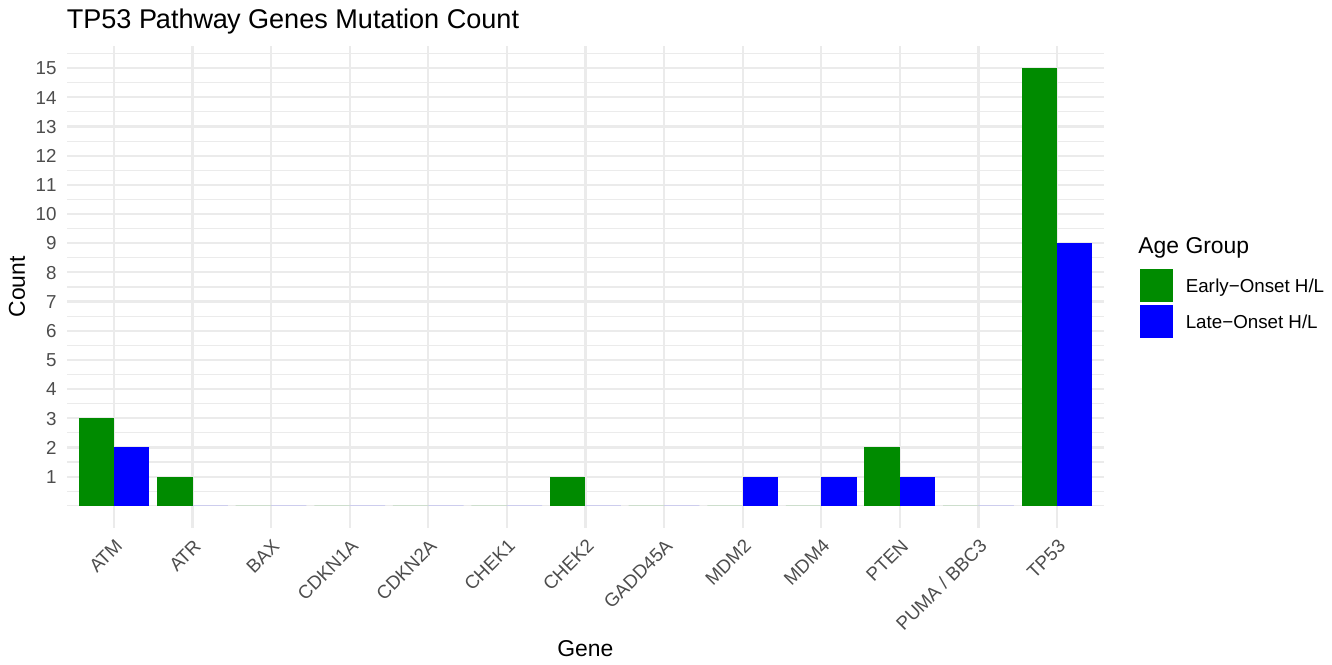

**Table S2.** Rates of PI3K and TP53 pathway alterations in early-onset and late-onset Hispanic/Latino CRC patients, stratified by colon and rectal adenocarcinomas.

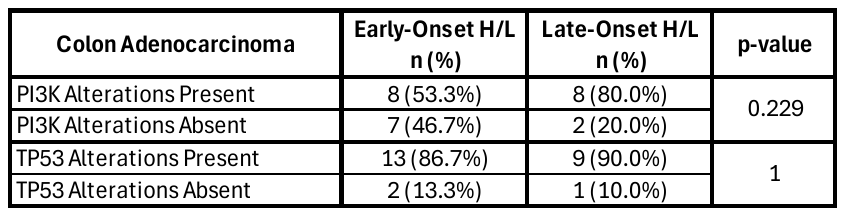

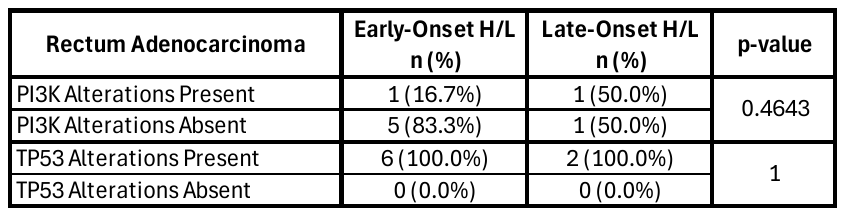

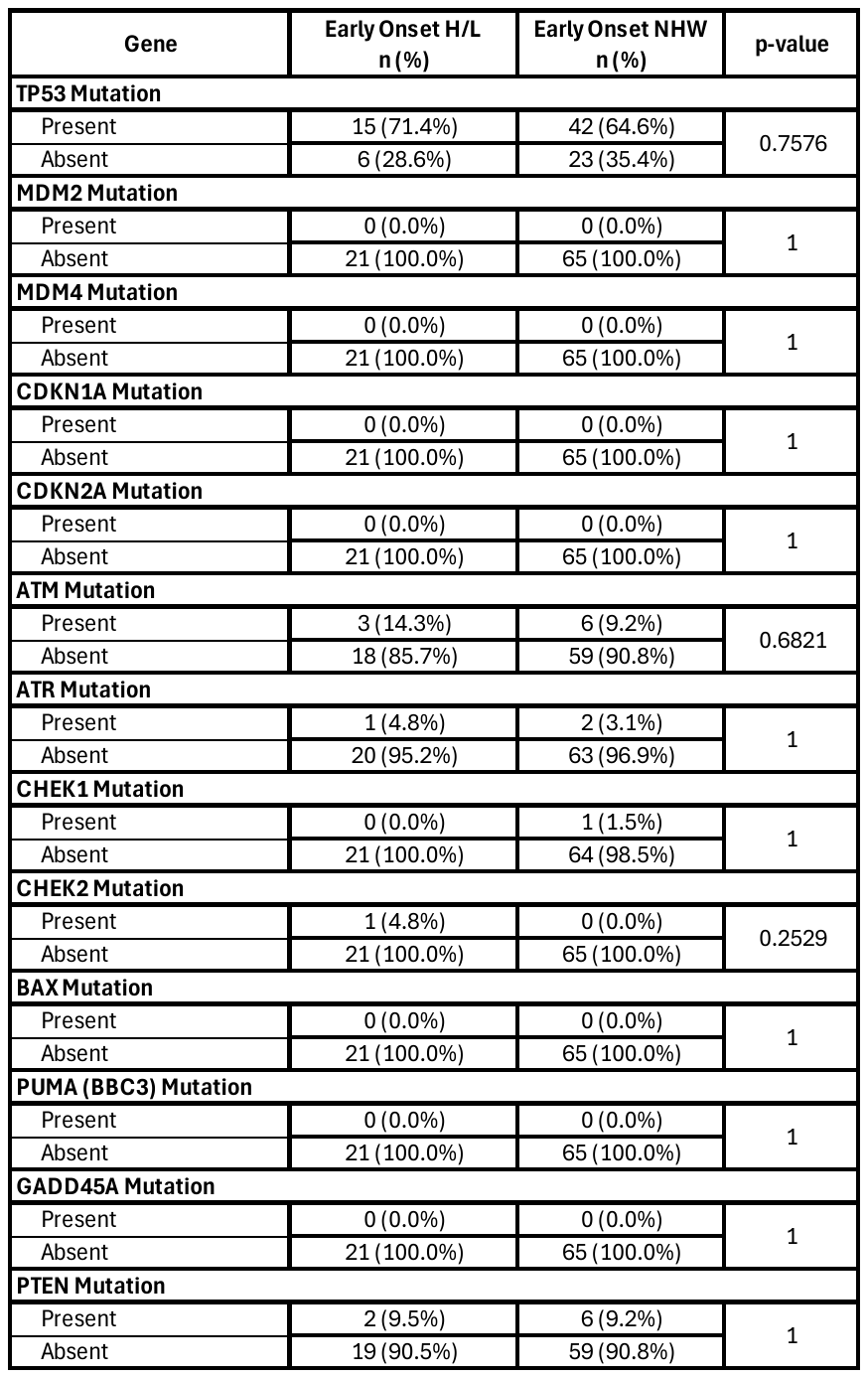
**Table S3.** Alteration Rates of PI3K and TP53 Pathway-Related Genes in Early-Onset Hispanic/Latino and Non-Hispanic White CRC Patients.

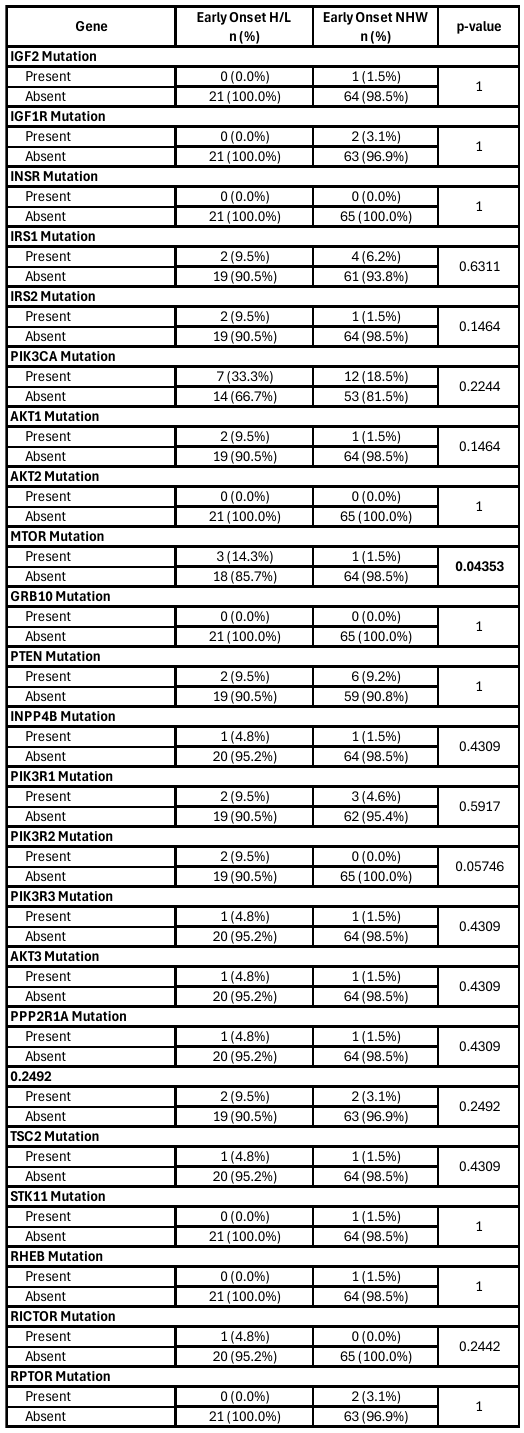

**Figure S3.** Counts of PI3K and TP53 Pathway-Related Gene Alterations in Early-Onset Hispanic/Latino and Non-Hispanic White CRC Patients.

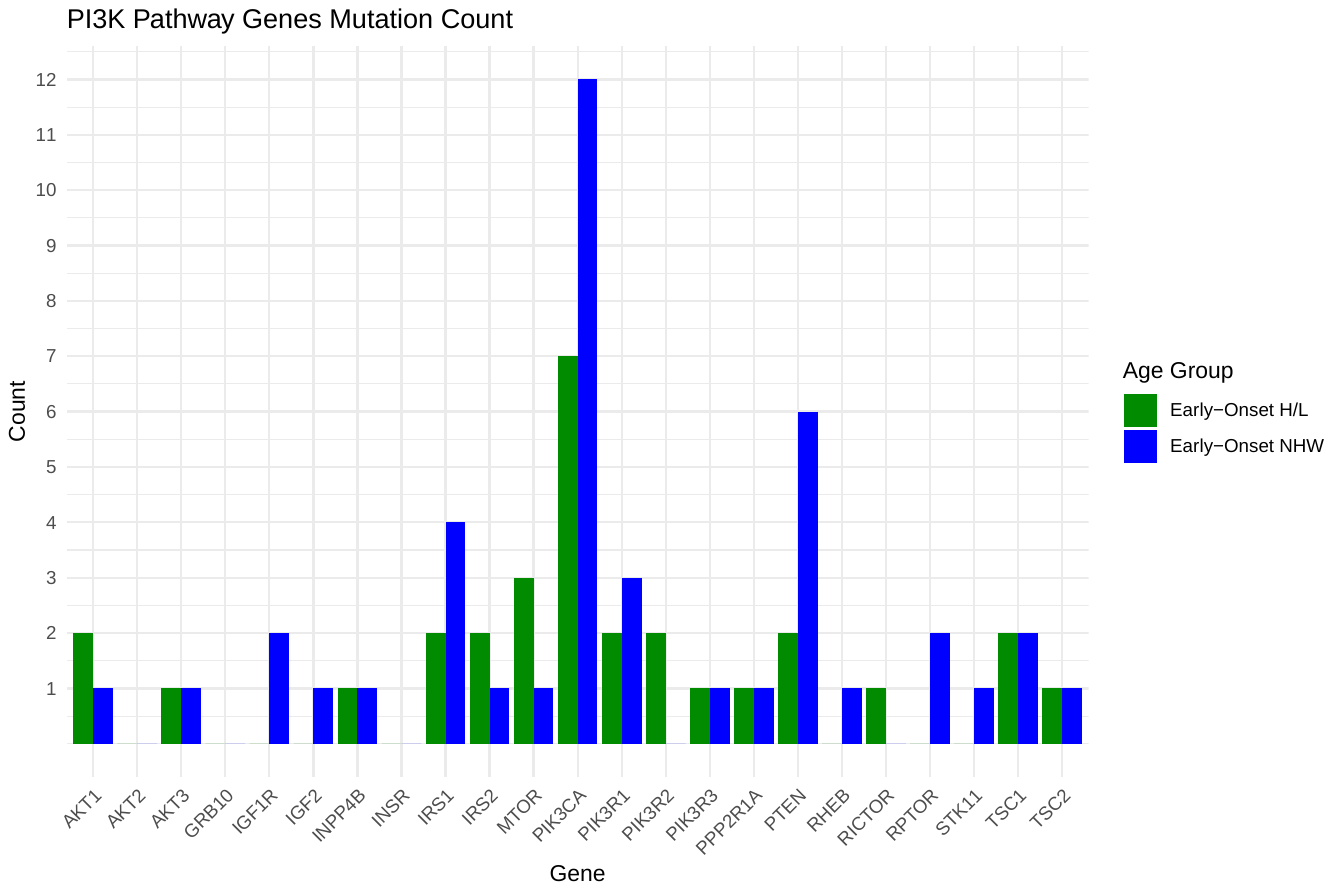

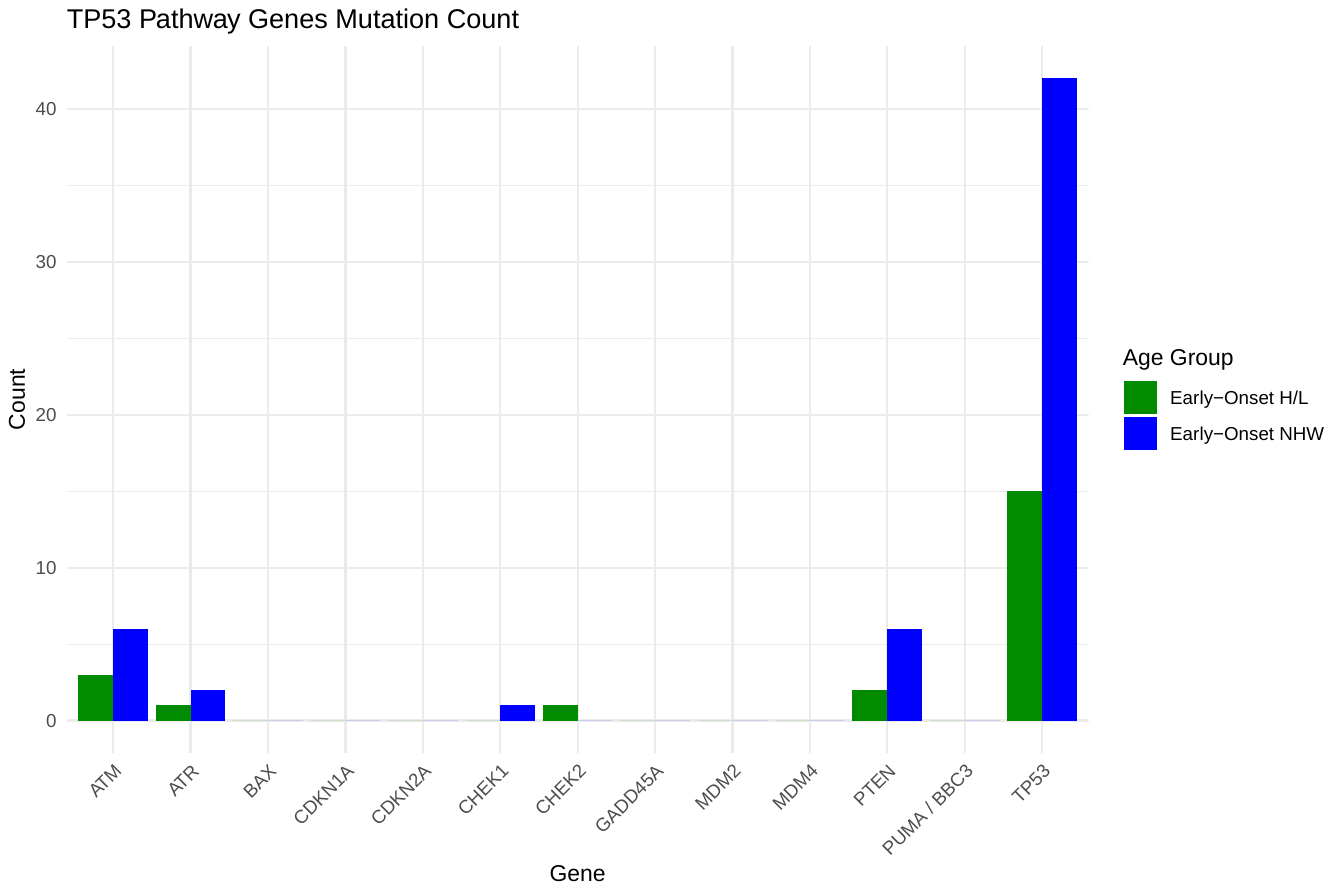

**Table S4.** Rates of PI3K and TP53 pathway alterations in early-onset Hispanic/Latino and Non-Hispanic White (NHW) CRC patients, stratified by colon and rectal adenocarcinomas.

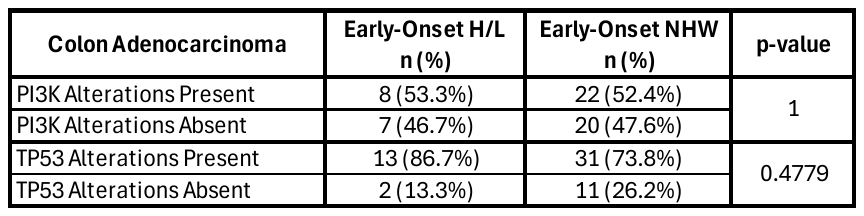

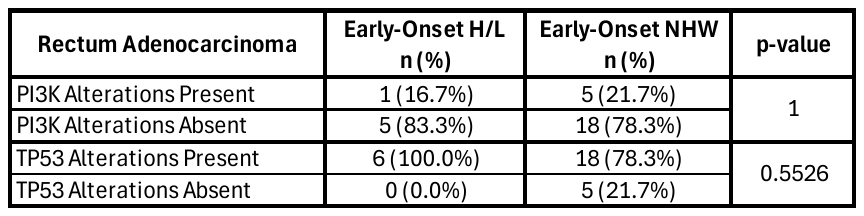
